# Impact of timing of RUTF dose reduction and visit frequency on acute malnutrition treatment effectiveness in Mali: A 2x2 Factorial Cluster-Randomized Controlled Trial

**DOI:** 10.64898/2026.01.26.26344816

**Authors:** Suvi T. Kangas, Zachary Tausanovitch, Césaire T. Ouédraogo, Issa N. Coulibaly, Christian Ritz, Bernardette Cichon, André Briend, Jeanette Bailey

## Abstract

**Background:** Severe acute malnutrition (SAM) among children under 5 years of age is generally treated in outpatient settings providing caregivers with weekly ready-to-use therapeutic food (RUTF) rations to be administered at home. Recent updates to global treatment guidelines suggest that RUTF dose can be reduced once children progress to moderate stage (MAM). No evidence exists on the optimal timing of the dosage reduction or on ideal visit frequency.

**Objectives:** We aimed to test the impact of 1) immediate RUTF dose reduction (1a) versus including a 2-week transition (1b) among children admitted with SAM and 2) fortnightly (2a) versus weekly (2b) visit frequency during MAM phase among children admitted with SAM and MAM.

**Methods and findings:** This prospective cluster-randomized controlled non-inferiority trial followed a 2 x 2 factorial design and divided 39 health areas (clusters) of Nara, Mali, into 4 groups implementing: A) 1a+2a B) 1a+2b, C) 1b+2a, and D) 1b+2b. Simplified, combined treatment was used providing 2 daily RUTF sachets to children admitted as SAM (mid-upper arm-circumference=MUAC<115mm or edema) and 1 daily RUTF sachet to children admitted with MAM (MUAC 115-124mm). Recovery was declared when a child reached MUAC ≥125mm and absence of edema for 2 consecutive visits. Depending on the randomization arm, children admitted with SAM transitioned into receiving 1 daily RUTF sachet immediately upon reaching MUAC≥115mm (A+B) or after 3 consecutive visits (2 weeks) with MUAC≥115mm (C+D). Weekly visits were applied for all children with MUAC<115mm and then depending on randomization arm, visits continued weekly (A+C) or fortnightly (B+D) in the MAM phase. The main outcome was recovery and a non-inferiority margin of 10% was applied. Between April and December 2023, a total of 6249 children with MUAC<125mm or edema were admitted to treatment including 1451 children with SAM. On average 98% of children recovered with a mean duration of treatment of 6 weeks. Immediate transition resulted in non-inferior recovery compared to 2-week transition from SAM to MAM and no differences were observed in program outcomes (proportion of recovered, defaulted, non-recovered, transferred to inpatient care and deceased). However, we observed a non-significant trend of slight increase in the proportion of children regressing back to SAM after being MAM among children following immediate transition compared to those benefitting from 2-week transition. Fortnightly visit frequency in MAM phase resulted in non-inferior recovery compared to weekly visits throughout and no differences were observed in program outcomes. Duration of treatment was 2.5 weeks longer with fortnightly visits during MAM phase resulting in 23 sachets higher RUTF consumption compared to weekly visits throughout.

**Conclusions:** We recommend applying weekly visits throughout treatment where feasible for both children with MAM and SAM at admission and including a 2-week transition period before reducing the RUTF dose for children admitted with SAM once they reach MAM criteria.

**Trial registration:** The study was registered to clinicaltrials.gov (NCT06594341).

## INTRODUCTION

In 2024, it was estimated that over 42.8 million infants and children between 6 months and five years of age experienced wasting at any given time [1]. Wasting dramatically increases the risk of death: compared to well-nourished children, children with moderate wasting (weight-for-height z-score (WHZ) between -3 and -2 standard deviation (SD)) are 3.4 times more likely to die, and this goes up to a staggering 11.6 times in children with severe wasting (WHZ < -3 SD)[2].

Treatment of wasting has, since 1981, been guided by WHO global recommendations that initially focused solely on inpatient treatment [3,4] but since 2007 also cover outpatient treatment of severe acute malnutrition (SAM) [5,6] and since 2023 include recommendations for the management of moderate acute malnutrition (MAM) [7]. Updates to WHO recommendations have wide implications to practice and even small adjustments to any aspects of treatment protocols can significantly impact effectiveness, uptake, coverage, cost and ultimately child survival.

One change in the 2023 guidelines is RUTF dose reduction once a child with SAM reaches the MAM phase. The evidence underlying the dose reduction recommendation [8] is based on 3 trials [9–11] which each had a different dose and reduction logic. Only the Maust et al. (2015) study tested reduction immediately upon reaching MAM while the Cazes et al. (2023) study reduced the dose at 2 anthropometric cut-off points (at mid-upper arm circumference = MUAC≥115mm and again at MUAC≥120mm) and the Kangas et al. (2019) study reduced the dose after 2 weeks into treatment regardless of anthropometry. Other studies not included in the WHO commissioned review have tested other dose reductions: the ComPAS trial tested a reduction only after 2 weeks with MAM criteria [12,13] while the OPTIMA+ComPAS trial in Niger [14,15] switched all children in both intervention arms immediately upon reaching MAM. Whether the RUTF reduction should be applied immediately once the child reaches MAM or after some weeks to ensure safe transition remains unclear with no guidance from WHO on the matter.

Another aspect of wasting treatment that has received less attention is the optimal visit frequency in outpatient care. Currently, there is no global guidance on recommended visit frequency. The practice in many national protocols is to recommend weekly visits for children with SAM and fortnightly visits for children with MAM. The impact of visit frequency on treatment effectiveness remains poorly documented. Only one study has tested this showing that monthly instead of weekly visits during SAM treatment resulted in 6.8% lower recovery [16]. However, the impact of a switch from weekly to fortnightly visit schedule remains unknown.

This study aimed to test two aspects of optimization to wasting treatment: 1) the timing of the reduction of RUTF dose during a SAM to MAM treatment transition and 2) the frequency of visits in the MAM phase of treatment.

## METHODS

### Study setting and population

The study was conducted in the catchment area of the Nara health district in southwestern Mali. The Nara region is characterized by a semi-arid tropical climate and an unstable security situation. These two situations exacerbate the already dire acute malnutrition situation, which was categorized as phase 3 (serious) by the Integrated Food Security Classification (IPC) [17] in 2023. In the same year, the prevalence of global acute malnutrition (GAM), the combination of SAM and MAM, according to WHZ (WHZ <-2) was 12.4% and SAM prevalence (WHZ <-3) was 2.0% [18].

At the beginning of the present study in April 2023, the Nara Health District was made up of 39 health areas, each comprising a community health center (CSCom=Centre de Sante Communautaire) as well as often one or more community health worker (CHW) sites offering malnutrition treatment. Since 2018, a simplified and combined protocol has been followed in the district to treat malnourished children at all treatment sites (CSComs and CHW sites) [19].

The simplified, combined treatment protocol admits children with MUAC<125mm or edema and treats those with MUAC<115mm or edema with 2 daily RUTF sachets while children with MUAC≥ 115mm get 1 daily RUTF sachet until discharge [12,13]. Since the switch to simplified, combined treatment in Nara, all children have been followed up weekly regardless of anthropometric status and children admitted with MUAC<115mm or edema have been switched to receive 1 daily RUTF sachet once they reach MUAC≥115mm and absence of edema upon 3 consecutive visits (=2 weeks minimum) [19].

### Study design

This was a prospective cluster-randomized controlled study following a 2x2 factorial design assessing the effect of 1) visit frequency during MAM phase (weekly vs. fortnightly) and 2) 2-week transition phase from SAM to MAM (included or suppressed), resulting in the following 4 randomization arms: A) fortnightly visit frequency for the MAM and elimination of the transition phase for SAM; B) weekly visit frequency for all children and elimination of the transition phase for SAM; C) fortnightly visit frequency for MAM and inclusion of a 2-week transition phase for SAM; and D) weekly visit for all children and inclusion of a 2-week transition phase for SAM (Supplementary Figures). The health area was defined as a cluster. A factorial design was selected to maximize power to detect effects for two factors assuming no interaction between the two factors. The study was registered to clinicaltrials.gov retrospectively on the 6^th^ of September 2023 (NCT06594341). The study protocol is included as Supplementary file 1 and a CONSORT check-list as Supplementary file 2.

### Randomization

The randomization of health areas was carried out in the presence of the district’s head physician. The project’s data manager prepared a list of the 39 health areas with their respective populations. He then ranked them in ascending order according to their populations and for the 4 largest health areas randomized them into the 4 arms using a computerized randomization (Microsoft VBA). The following 24 health areas were then randomized in blocs of 12 and the final 11 health areas were randomized as a final bloc. After randomization, the district’s head physician communicated the list of centers and the arm to which they belonged to each health center director. Study teams then visited the centers to support them in the implementation of the changes to the treatment protocol.

### Outcomes

The primary outcome was recovery. Secondary outcomes included defaulting, non-recovery, transfer to inpatient care and deceased, regressed to SAM after being MAM, as well as duration of treatment until discharge, RUTF consumption, number of visits observed, number of missed visits, and MUAC and weight gain velocity from admission to discharge.

### Definitions

Recovery was defined as a MUAC≥125 mm and the absence of edema and medical complications during two consecutive visits. Non-recovery was defined as not having recovered by 16 weeks of treatment and defaulting as missing two consecutive visits. The recovery, defaulting, non-recovered, and deceased percentages were calculated over these four exit categories as per the community-based management of acute malnutrition reporting guidance [20]. Transfers to inpatient care are reported but not included in the programmatic outcome denominator. Early recoveries (children discharged recovered while only reaching recovery criteria once) and unknown outcomes were classified to defaulters.

Duration of treatment was calculated as the days from admission to outpatient program until discharge, with discharge being the last visit the child was cared for at the treatment site. Per standard and recommended practice [21], weight gain velocity was calculated (Kamugisha et al., 2021) in terms of relative increase per day as the discharge weight minus minimum weight in grams divided by the minimum weight in kilos and divided by the days from minimum weight to discharge. Per usual practice [22], MUAC gain velocity was calculated in terms of absolute increase per week as the discharge MUAC minus minimum MUAC in mm divided by the weeks from minimum MUAC to discharge.

The number of visits observed was calculated as the total number of treatment visits the child attended. Conversely, the number of missed visits was calculated as the total number of visits the child missed, excluding any visits that were planned to be skipped and that benefitted from double RUTF rations.

The amount of RUTF consumed per child was calculated as the number of sachets prescribed during the treatment including the 7 sachets exit ration given at discharge.

### Sample size estimations

The sample size was calculated to demonstrate two hypotheses: 1) a non-inferior recovery in children admitted with MUAC<125mm or edema when following fortnightly visits in the MAM phase compared to when applying weekly visits; and 2) a non-inferior recovery in children with MUAC<115mm or edema at admission when removing the 2-week SAM-MAM transition phase compared to when maintaining the 2-week transition phase. As the second hypothesis involves a sub-sample (MUAC<115mm or edema) of the first (MUAC<125mm or edema), the second hypothesis was used as the basis of the sample size calculation.

With a fixed number of clusters (n=39) and combining 2 randomization arms each time to analyze the differences, this results in 19-20 clusters combined per intervention arm. Using a minimum of 19 clusters per arm, a maximum difference of 3 percentage points in proportion recovered between control and intervention, a 10% non-inferiority margin, power of 80%, a probability of type 1 error of 2.5%, and an intra-cluster correlation coefficient of 0.05, a sample size of 40 children were required per cluster resulting in a total of 1,560 children with MUAC<115mm and/or edema to be included in the study. Assuming children with MUAC<115mm or edema represent 35% [19], a total of 4680 children with MUAC<125mm or edema were expected to be included in total.

### Data collection

Routine data on malnourished children admitted to treatment was collected initially by routine health staff on paper-based registries and individual cards as per the Mali national protocol (annexes 16 and 18 in the National protocol) [23]. Select data including visit dates, anthropometric measurements and prescription of RUTF at admission and each follow-up visit until discharge was then collected on electronic forms retrospectively during monthly visits by study staff to the treatment sites.

### Data analysis

Descriptive statistics are presented by randomization arm as percentages and frequencies for categorical variables and means with standard deviations (SD) and median with interquartile range (IQR) for continuous variables that were normally and non-normally distributed, respectively.

The impact of spacing out follow-up visits and eliminating the transition phase on the treatment effectiveness was assessed using linear and logistic mixed models with treatment sites and health areas (clusters) as random effects. Impact of visit frequency spacing was estimated for all children admitted (GAM), for those with MAM at admission and those with SAM at admission, while impact of transition phase was only estimated for children with SAM at admission. Non-inferiority of recovery was assessed by means of a 97.5% one-sided confidence interval (assessing whether or not the interval was within the non-inferiority margins) and visualized using a 95% two-sided confidence interval. For other outcomes, estimates and 95% two-sided confidence intervals were reported. Mean differences between interventions were estimated by combining groups A, B, C and D, and comparing A+B to C+D for assessing the transition effect, and comparing A+C to B+D for assessing the effect of visit spacing.

Unadjusted models and models adjusted for sex, age, admission height, MUAC, WHZ, height-for-age (HAZ) and month were fitted. Collinearity between adjustment variables was checked and correlations <0.6 allowed. If models didn’t converge, random effects were dropped one by one (first treatment site, then health area). All models included the adjustment for the other factor (in case of estimating impact of visit frequency on outcomes, transition variable was included in adjustments and vice versa).

All analyses were conducted applying both the “de facto” principle of real life results that we call intention to treat (ITT) and “de juro” principle of best case scenario that we call per protocol (PP) for the main outcome and the secondary outcomes [24]. The ITT analysis included all children admitted to the study as originally allocated after randomization and for whom outcome data is available. The PP analysis included all children but accounted for protocol violation. Protocol violations included anyone who obtained wrong RUTF dose or who missed visits (excluding scheduled missed visits that benefitted from double RUTF ratios).

To explore sub-group effects, linear and logistic mixed models with two-way subgroup-treatment interaction terms were fitted . Potential subgroups were sex (male/female), age (<12 months/≥12 months), MUAC (<115 mm/ ≥115 mm), WHZ (<-3/≥-3), HAZ (<-2/≥-2) and the other factor (weekly vs fortnightly visit frequency in case of analysis of transition and vice versa). Data were analyzed using STATA version 17 [25]. A p-value < 0.05 was considered statistically significant. Intracluster correlation coefficients (ICC) for health area and treatment site were reported from the unadjusted models on recovery and duration of treatment reporting the random effects coefficients.

### Ethical approval

Ethical approvals for the study were obtained from the ethics committee of the Faculté de Médicine et d’Odonto-stomatologie de l’Université des Sciences Techniques et Technologies de Bamako in Mali (N°2023/38/CE/USTTB) and the IRC institutional review board (H1.00.025).

No individual consent was requested from participants since the primary data collection was based on routine data and the secondary data collection for research purposes happened retrospectively based on routine data. For research inclusivity purposes [26], a French version of the manuscript is provided as Supplementary File 3. The database used in the current work is available from the Zenodo data repository (https://doi.org/10.5281/zenodo.17716792).

## RESULTS

### Participant profile

In total, 6249 children with MUAC<125mm or edema were admitted between April and December 2023 to the 39 health areas randomized to one of the four arms, including 1452 children with MUAC<115mm or edema. Total number admitted per arm ranged from 1761 to 1458 for GAM and 291 to 430 for SAM.

**Figure 1:**
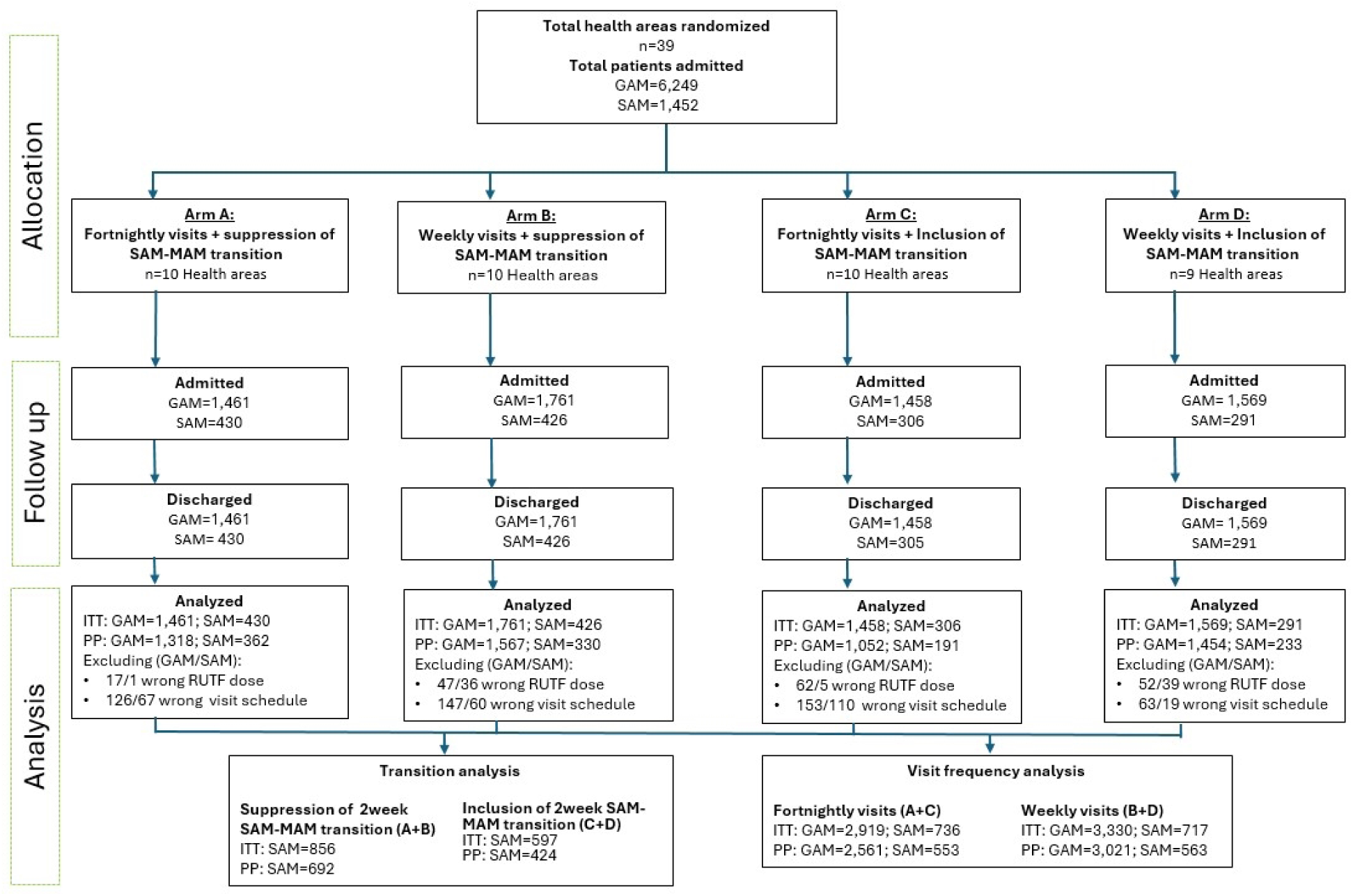
Randomization of health areas and inclusion of participants in the study on the impact of timing of RUTF dose reduction and visit frequency on acute malnutrition treatment effectiveness in Mali

Children were on average 15 months of age at admission, 45% were male, their median [IQR] MUAC was 119 [115;120] mm and weight 6.7 [6.0; 7.5] kg at admission to treatment (Table 1). No differences in anthropometry were observed between arms at admission to treatment (Supplementary Table 1). There was only 1 child with edema in group C. Most (98% of) children recovered during treatment with 40 cases of early recovery and 20 unknown outcomes classified to defaulters.

**Table 1:** Characteristics of children at admission to treatment according to randomization arm and by severity of acute malnutrition (GAM, SAM, MAM)

|  | **A:Fortnightly visits + suppression of SAM-MAM transition** | **B:Weekly visits + suppression of SAM-MAM transition** | **C:Fortnightly visits + SAM-MAM transition** | **D:Weekly visits + SAM-MAM transition** |
| --- | --- | --- | --- | --- |
| **GAM, % (n)** | **100.0 (1461)** | **100.0 (1761)** | **100.0 (1458)** | **100.0 (1569)** |
| Age, median[IQR] | 13.0 [10.0;21.0] | 13.0 [10.0;20.0] | 13.0 [9.0;20.0] | 13.0 [10.0;18.0] |
| Male, n (%) | 43.5 (635) | 45.8 (806) | 45.4 (662) | 45.8 (718) |
| Edema, % (n) | 0.0 (0) | 0.0 (0) | 0.1 (1) | 0.0 (0) |
| MUAC (mm), median [IQR] | 118.0 [113.0;120.0] | 119.0 [115.0;120.0] | 118.0 [115.0;120.0] | 120.0 [116.0;122.0] |
| weight (kg), median [IQR] | 6.7 [6.0;7.5] | 6.7 [6.0;7.5] | 6.7 [5.9;7.5] | 6.7 [6.0;7.3] |
| height (cm), median [IQR] | 72.0 [68.0;76.0] | 71.2 [67.0;76.0] | 70.6 [66.5;75.2] | 71.0 [67.5;75.5] |
| WAZ (sd), median [IQR] | -3.1 [-3.9;-2.5] | -3.2 [-4.0;-2.4] | -3.2 [-4.1;-2.5] | -3.2 [-4.0;-2.4] |
| HAZ (sd), median [IQR] | -2.0 [-3.0;-1.0] | -2.0 [-3.1;-0.9] | -2.1 [-3.3;-0.9] | -1.9 [-2.9;-0.8] |
| WHZ (sd), median [IQR] | -2.86 ± 0.04 | -2.94 ± 0.04 | -2.74 ± 0.05 | -3.12 ± 0.04 |
| **MAM, % (n)** | **70.6 (1031)** | **75.8 (1335)** | **79.0 (1152)** | **81.5 (1278)** |
| Age, median[IQR] | 14.0 [10.0;22.0] | 13.0 [10.0;20.0] | 13.0 [10.0;22.0] | 13.0 [10.0;18.0] |
| Male, n (%) | 43.4 (447) | 46.8 (625) | 46.4 (534) | 46.6 (595) |
| MUAC (mm), median [IQR] | 0.0 (0) | 0.0 (0) | 0.0 (0) | 0.0 (0) |
| weight (kg), median [IQR] | 120.0 [117.0;121.0] | 120.0 [118.0;121.0] | 119.0 [117.0;121.0] | 120.0 [118.0;122.0] |
| height (cm), median [IQR] | 7.0 [6.3;7.7] | 6.9 [6.2;7.7] | 6.9 [6.1;7.7] | 6.8 [6.2;7.4] |
| WAZ (sd), median [IQR] | 72.2 [68.5;76.5] | 72.0 [68.2;76.2] | 71.5 [67.5;76.0] | 71.5 [68.4;76.1] |
| HAZ (sd), median [IQR] | -2.9 [-3.5;-2.2] | -3.0 [-3.8;-2.3] | -3.0 [-3.8;-2.3] | -3.1 [-3.8;-2.3] |
| WHZ (sd), median [IQR] | -1.8 [-2.8;-0.9] | -1.9 [-2.8;-0.8] | -1.9 [-3.1;-0.8] | -1.8 [-2.8;-0.7] |
| **SAM, % (n)** | **29.4 (430)** | **24.2 (426)** | **21.0 (306)** | **18.6 (291)** |
| Age, median [IQR] | 13.0 [10.0;19.0] | 12.0 [9.0;18.0] | 12.0 [8.0;16.0] | 12.0 [9.0;18.0] |
| Male, n(%) | 43.7 (188) | 42.5 (181) | 41.8 (128) | 42.3 (123) |
| Edema, % (n) | 0.0 (0) | 0.0 (0) | 0.3 (1) | 0.0 (0) |
| MUAC (mm), median [IQR] | 110.0 [108.0;113.0] | 110.0 [108.0;113.0] | 110.0 [102.0;113.0] | 111.0 [109.0;113.0] |
| weight (kg), median [IQR] | 6.1 [5.5;6.8] | 5.8 [5.2;6.6] | 5.7 [5.0;6.4] | 5.9 [5.2;6.6] |
| height (cm), median [IQR] | 70.3 [67.0;74.5] | 68.4 [64.6;73.8] | 68.0 [64.0;72.0] | 68.5 [64.3;72.8] |
| WAZ (sd), median [IQR] | -3.9 [-4.6;-3.1] | -4.0 [-4.8;-3.4] | -4.1 [-4.7;-3.4] | -3.9 [-4.8;-3.2] |
| HAZ (sd), median [IQR] | -2.3 [-3.8;-1.2] | -2.5 [-3.8;-1.5] | -2.6 [-3.8;-1.6] | -2.5 [-3.8;-1.4] |
| WHZ (sd), median [IQR] | -3.6 [-4.2;-2.8] | -3.5 [-4.4;-2.7] | -3.5 [-4.2;-2.9] | -3.6 [-4.7;-2.8] |
| Abbreviations: GAM, global acute malnutrition (SAM+MAM); HAZ, height-for-age z-score; MAM, moderate acute malnutrition; MUAC, mid-upper arm circumference; SAM, severe acute malnutrition; WAZ, weight-for-age z-score; WHZ, weight-for-height z-score. |  |  |  |  |

### Impact of visit frequency in MAM phase

The proportion of children recovered following treatment with fortnightly visits was non-inferior to the proportion of children recovered following weekly visits (p-value for non-inferiority) both in ITT (p<0.001) and in PP (p<0.001) (Figure 2). No differences were observed in the proportion of children recovered, defaulted, non-recovered, transferred or deceased by visit frequency (Table 2). In unadjusted analyses, children following weekly visits had 2.8 weeks [95%CI: 2.4 to 3.2] shorter duration of treatment, 1.0 g/kg/d [95%CI: 0.6, 1.3] higher weight gain velocity and

**Figure 2:**
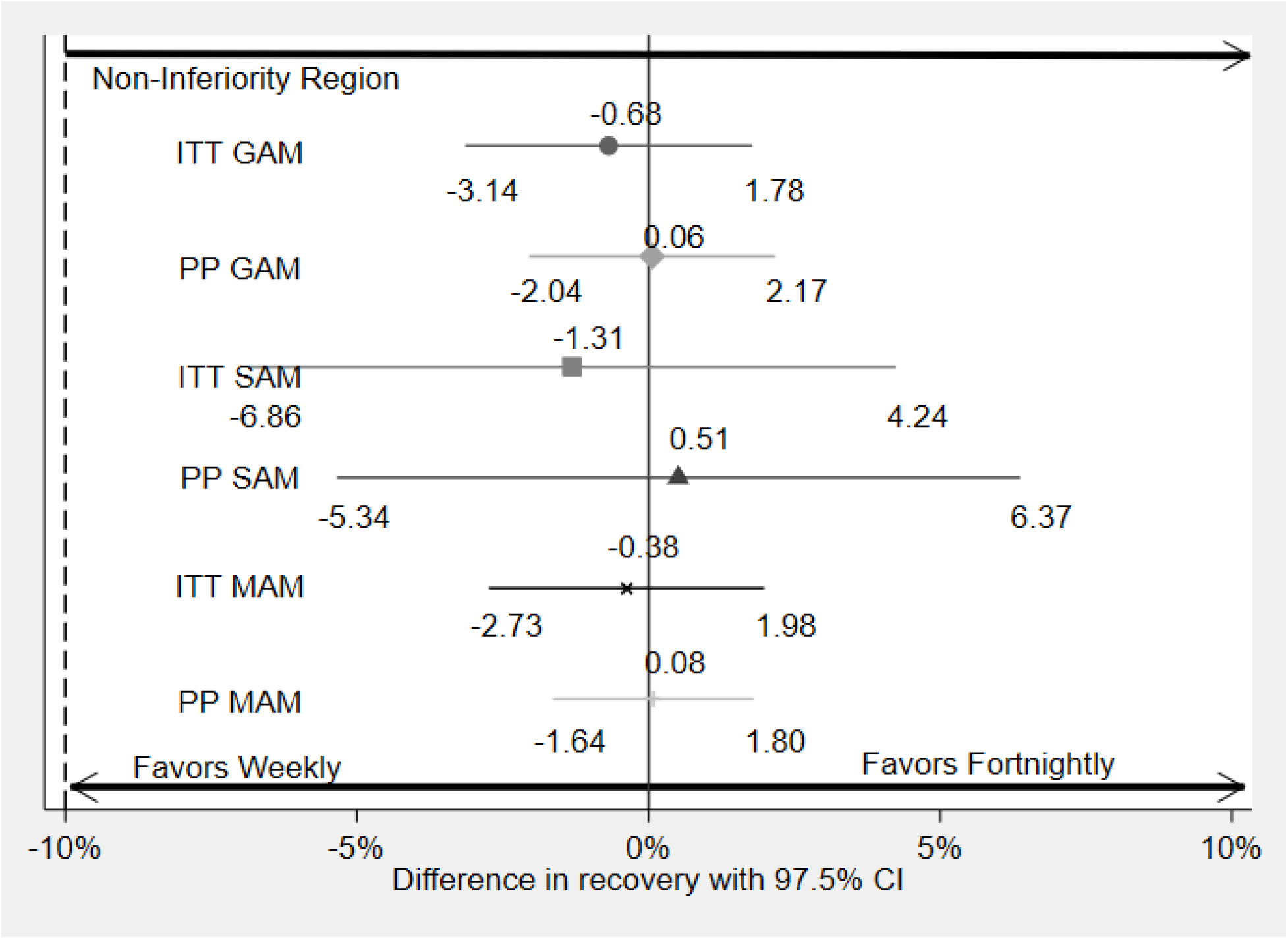
Non-inferiority of recovery with fortnightly visit frequency compared to weekly visit frequency in the MAM phase of treatment in adjusted analysis. Abbreviations: GAM, global acute malnutrition (SAM+MAM); ITT, intention to treat; MAM, moderate acute malnutrition; PP, per protocol; SAM, severe acute malnutrition.

**Table 2:** Impact of visit frequency in MAM phase on treatment outcomes among children with MUAC<125mm or edema admitted to simplified treatment in intention-to-treat analysis

| Treatment outcomes | Visit frequency in MAM phase |  | Difference [95%CI] |  |
| --- | --- | --- | --- | --- |
|  | Weekly | Fortnightly | Unadjusted | Adjusted* |
| **GAM, n** | 3330 | 2919 |  |  |
| **Primary outcome** |  |  |  |  |
| Recovered, % (n) | 98% (3260) | 98% (2834) | -0.73% [-2.71%, 1.24%] | -0.68% [-3.14%, 1.78%] |
| **Secondary outcomes** |  |  |  |  |
| Defaulted, % (n) | 1.7% (56) | 1.8% (53) | -0.02% [-1.59%, 1.55%] | 0.16% [-1.87%, 2.20%] |
| Non-recovered, % (n) | 0.3% (9) | 0.9% (25) | 0.59% [-0.16%, 1.35%] | 0.86% [-0.04%, 1.76%] |
| Transferred, % (n) | 0.1% (4) | 0.2% (7) | 0.21% [-153.74%, 154.16%] | 0.07% [-1.48%, 1.62%] |
| Died, % (n) | 0.03% (1) | 0.0% (0) | -0.03% [-0.09%, 0.03%] |  |
| Developed SAM after being MAM, % (n) | 0.8% (27) | 0.7% (20) | -0.01% [-0.80%, 0.77%] | 0.06% [-0.89%, 1.01%] |
| Duration of treatment (weeks), median [IQR] | 4 [3;6] | 8 [6;10] | 2.77 [2.35, 3.20] | 2.49 [2.04, 2.94] |
| Weight gain velocity (g/kg/d), mean ± SD | 4.9 ± 2.3 | 4.0 ± 1.9 | -0.95 [-1.29, -0.60] | -0.86 [-1.36, -0.36] |
| MUAC gain velocity (mm/w), mean ± SD | 2.2 ± 0.6 | 1.6 ± 0.6 | -0.54 [-0.70, -0.39] | -0.58 [-0.78, -0.39] |
| Number of treatment visits, median [IQR] | 5 [4;7] | 5 [4;6] | -0.62 [-0.95, -0.29] | -0.84 [-1.15, -0.53] |
| Number of missed visits, median [IQR] | 0 [0;0] | 0 [0;0] |  | 0.03 [-0.02, 0.07] |
| Amount of RUTF per child, median [IQR] | 35 [28;56] | 70 [56;84] | 26.61 [22.45, 30.78] | 22.87 [19.09, 26.66] |
| **MAM, n** | 2613 | 2183 |  |  |
| Recovered, % (n) | 99% (2584) | 98% (2148) | -0.25% [-1.87%, 1.37%] | -0.38% [-2.73%, 1.98%] |
| Defaulted, % (n) | 1.1% (29) | 1.3% (28) | 0.02% [-1.49%, 1.53%] | 0.10% [-1.82%, 2.01%] |
| Non-recovered, % (n) | 0% (0) | 0.2% (5) | 0.23% [0.03%, 0.43%] |  |
| Transferred, % (n) | 0% (0) | 0.1% (2) | 0.09% [-0.04%, 0.22%] |  |
| Died, % (n) | 0% (0) | 0% (0) |  |  |
| Developed SAM after being MAM, % (n) | 0.5% (14) | 0.4% (9) | -0.08% [-0.84%, 0.67%] | 0.18% [-0.96%, 1.31%] |
| Duration of treatment (weeks), median [IQR] | 4 [3;5] | 6 [6;8] | 2.70 [2.35, 3.05] | 2.56 [2.14, 2.97] |
| Weight gain velocity (g/kg/d), mean ± SD | 4.7 ± 2.2 | 3.6 ± 1.7 | -1.07 [-1.38, -0.76] | -0.90 [-1.38, -0.41] |
| MUAC gain velocity (mm/w), mean ± SD | 2.1 ± 0.6 | 1.5 ± 0.5 | -0.61 [-0.78, -0.44] | -0.66 [-0.87, -0.45] |
| Number of treatment visits, median [IQR] | 5 [4;6] | 4 [4;5] | -0.70 [-0.93, -0.48] | -0.78 [-1.07, -0.49] |
| Number of missed visits, median [IQR] | 0 [0;0] | 0 [0;0] | -0.02 [-0.05, 0.01] | -0.01 [-0.04, 0.02] |
| Amount of RUTF per child, median [IQR] | 35 [28;42] | 56 [56;70] | 25.40 [22.85, 27.94] | 23.82 [20.41, 27.24] |
| **SAM, n** | **717** | **736** |  |  |
| Recovered, % (n) | 94% (676) | 93% (686) | -1.21% [-4.87%, 2.45%] | -1.31% [-6.86%, 4.24%] |
| Defaulted, % (n) | 3.7% (27) | 3.4% (25) | -0.66% [-92.79%, 91.48%] | 0.13% [-4.10%, 4.35%] |
| Non-recovered, % (n) | 1.3% (9) | 2.7% (20) | 1.91% [-0.77%, 4.59%] | 3.66% [-0.21%, 7.52%] |
| Transferred, % (n) | 0.6% (4) | 0.7% (5) | 0.14% [-0.74%, 1.02%] | -1.13% [-5.05%, 2.79%] |
| Died, % (n) | 0.1% (1) | 0% (0) | -0.14% [-0.41%, 0.13%] |  |
| Developed SAM after being MAM, % (n) | 1.8% (13) | 1.5% (11) | -0.20% [-1.69%, 1.28%] | -0.75% [-3.10%, 1.60%] |
| Duration of treatment (weeks), median [IQR] | 8 [7;9] | 10 [8;12] | 2.39 [1.74, 3.04] | 2.09 [1.31, 2.88] |
| Weight gain velocity (g/kg/d), mean ± SD | 5.6 ± 2.3 | 5.0 ± 2.2 | -0.69 [-1.16, -0.22] | -0.63 [-1.29, 0.03] |
| MUAC gain velocity (mm/w), mean ± SD | 2.3 ± 0.6 | 2.0 ± 0.6 | -0.34 [-0.49, -0.20] | -0.34 [-0.53, -0.16] |
| Number of treatment visits, median [IQR] | 9 [7;10] | 8 [6;9] | -0.89 [-1.45, -0.33] | -1.10 [-1.65, -0.55] |
| Number of missed visits, median [IQR] | 0 [0;0] | 0 [0;0] | 0.22 [0.05, 0.40] | -0.01 [-0.04, 0.02] |
| Amount of RUTF per child, median [IQR] | 84 [70;105] | 112 [84;126] | 23.06 [16.89, 29.24] | 19.12 [12.71, 25.53] |
| * adjusted for sex, age, height, MUAC, weight-for-height z-score, height-for-age z-score, month of admission and transition (included or suppressed).<br>Abbreviations: GAM, global acute malnutrition (SAM+MAM); MAM, moderate acute malnutrition; MUAC, mid-upper arm circumference; RUTF, ready-to-use therapeutic food; SAM, severe acute malnutrition. |  |  |  |  |

26.6 sachets [95%CI: 22.5, 30.8] lower RUTF consumption compared to children following fortnightly visits in the MAM phase. When adjusting for admission characteristics, these differences remained unaltered. Similar results were observed when only looking at children admitted as MAM and SAM (Table 2), in PP (Supplementary Table 2) and when only looking at recovered children (Supplementary Table 3). No differences were observed in subgroup analyses (Supplementary tables 4-5). The ICC of recovery within a health area was 0.237 and for treatment site within a health area was 0.161 while ICC for duration of treatment was 0.112 for health area and 0.017 for treatment site within a health area.

### Impact of immediate transition from SAM to MAM among children with SAM

The proportion of children recovered following the immediate transition from SAM to MAM was non-inferior to the proportion of children recovered following the 2-week transition period (p-value for non-inferiority) both in ITT (p<0.001) and in PP (p<0.001) (Figure 3). No differences were observed in the proportion of children recovered, defaulted, non-recovered, transferred or deceased by the immediate or 2-week SAM to MAM transition (Table 3). In unadjusted analyses, children who had transitioned immediately once they reached MAM had a 0.9 g/kg/d [95%CI: 0.4, 1.4] lower weight gain velocity and 9.1 sachets [95%CI: 2.9, 15.3] lower RUTF consumption compared to children who waited for 2 weeks before transitioning to the MAM treatment phase. When adjusting for admission characteristics, the difference in weight gain velocity was no longer significant (mean Δ=0.65 [95%CI:-0.01; 1.32] g/kg/d) but the difference in RUTF persists. Immediate transition also seemed to result in a slight increase in the proportion of children regressing back to SAM after having been MAM although this difference was not statistically significant (mean Δ=1.79% [95%CI: -0.26%, 3.83%] in unadjusted analysis and mean Δ=3.17% [95%CI:-0.11%, 6.45%] in adjusted analysis). Similar results were observed among recovered only (Table 3) and in per protocol analyses (Supplementary Table 6). No significant effect modification of the intervention (by sex, age, WHZ category, HAZ category or visit frequency in MAM phase) on recovery or duration of treatment was observed (Supplementary tables 7-8).

**Figure 3:**
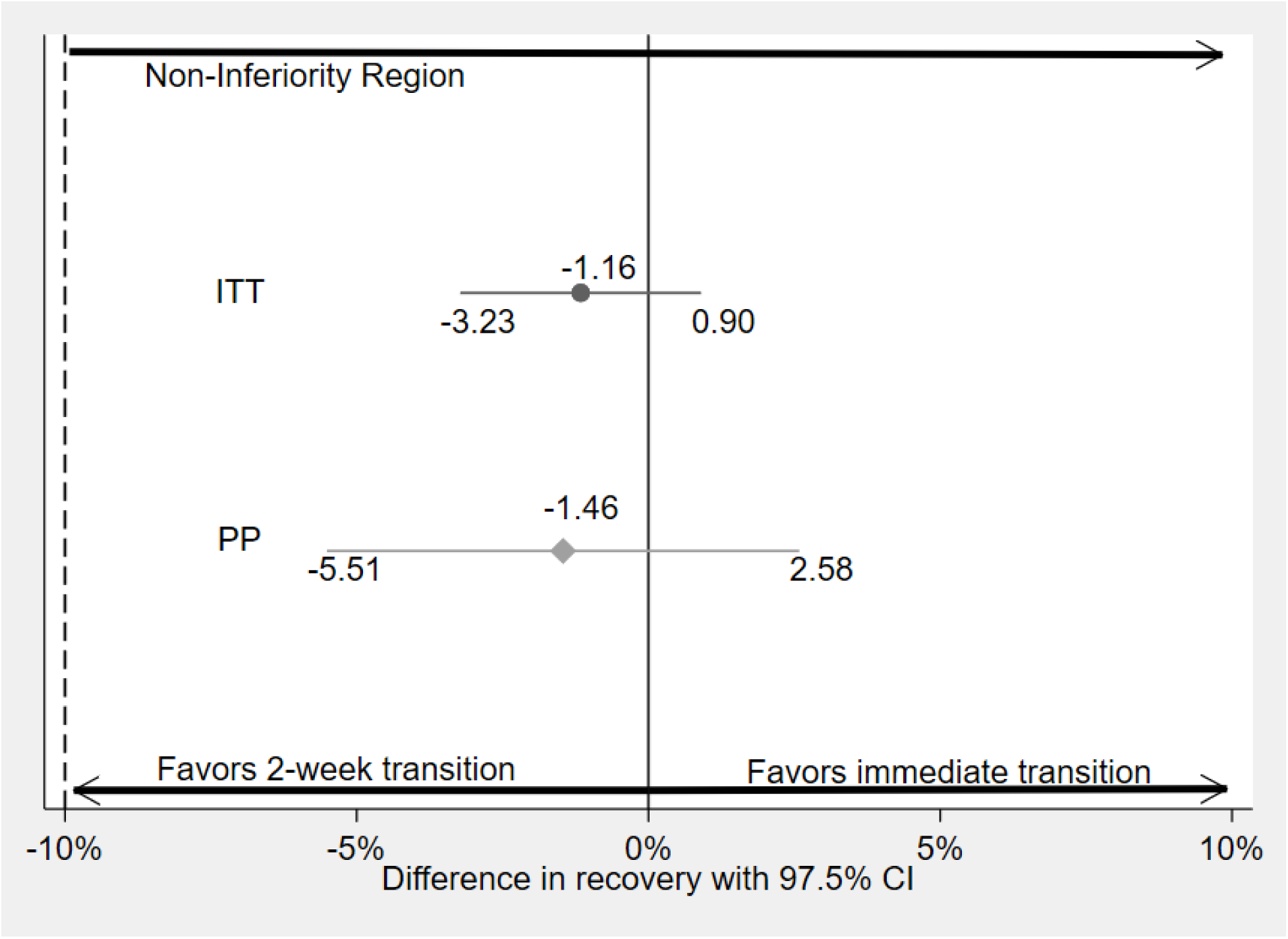
Non-inferiority of recovery following immediate transition from SAM to MAM compared to including the 2-week transition among children admitted with SAM in adjusted analysis. Abbreviations: ITT, intention to treat; MAM, moderate acute malnutrition; PP, per protocol; SAM, severe acute malnutrition.

**Table 3:**
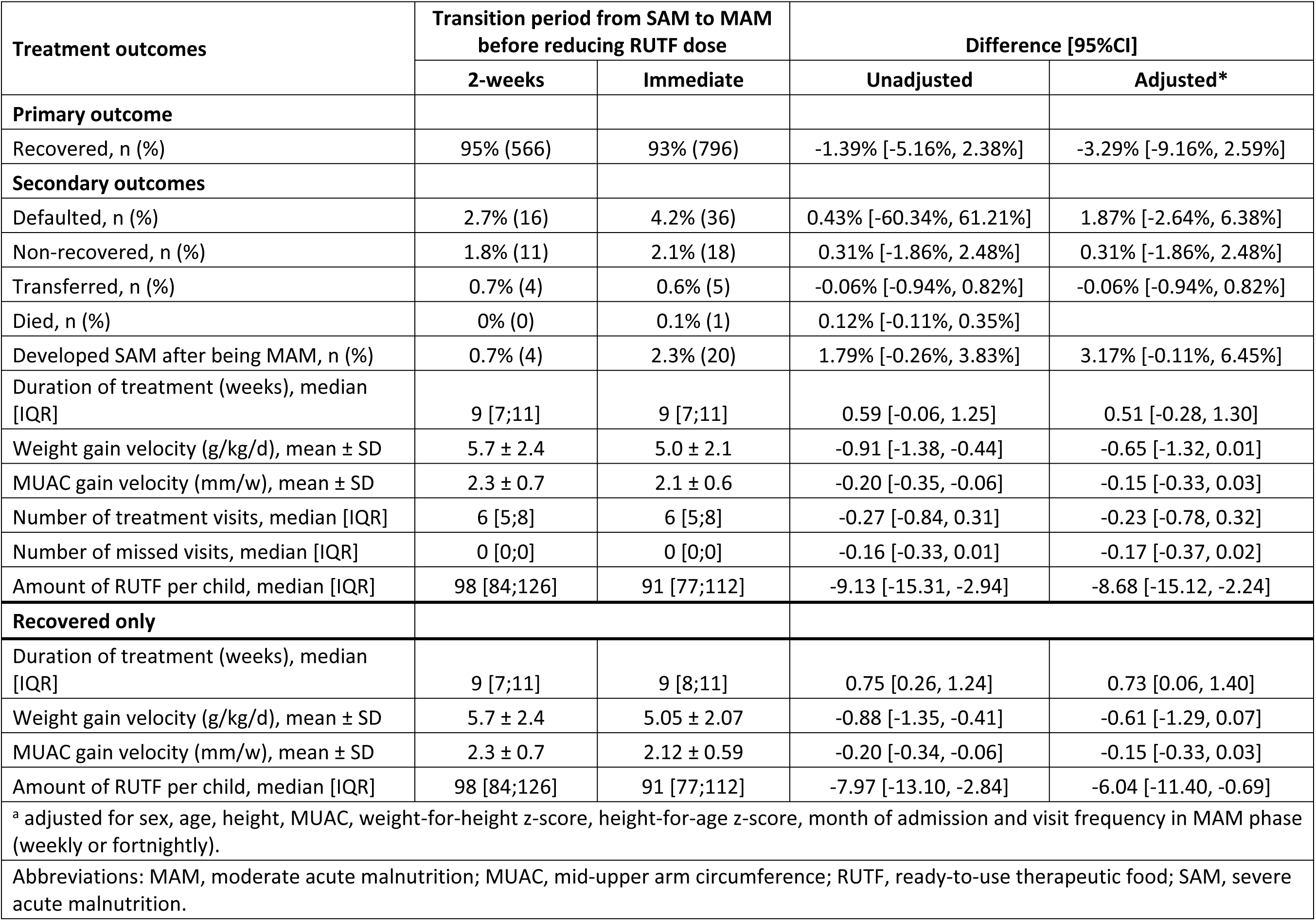
Impact of immediate SAM to MAM transition phase among children with MUAC<115mm or edema admitted to simplified treatment in intention-to-treat analysis

| Treatment outcomes | Transition period from SAM to MAM<br>before reducing RUTF dose |  | Difference [95%CI] |  |
| --- | --- | --- | --- | --- |
|  | 2-weeks | Immediate | Unadjusted | Adjusted* |
| **Primary outcome** |  |  |  |  |
| Recovered, n (%) | 95% (566) | 93% (796) | -1.39% [-5.16%, 2.38%] | -3.29% [-9.16%, 2.59%] |
| **Secondary outcomes** |  |  |  |  |
| Defaulted, n (%) | 2.7% (16) | 4.2% (36) | 0.43% [-60.34%, 61.21%] | 1.87% [-2.64%, 6.38%] |
| Non-recovered, n (%) | 1.8% (11) | 2.1% (18) | 0.31% [-1.86%, 2.48%] | 0.31% [-1.86%, 2.48%] |
| Transferred, n (%) | 0.7% (4) | 0.6% (5) | -0.06% [-0.94%, 0.82%] | -0.06% [-0.94%, 0.82%] |
| Died, n (%) | 0% (0) | 0.1% (1) | 0.12% [-0.11%, 0.35%] |  |
| Developed SAM after being MAM, n (%) | 0.7% (4) | 2.3% (20) | 1.79% [-0.26%, 3.83%] | 3.17% [-0.11%, 6.45%] |
| Duration of treatment (weeks), median [IQR] | 9 [7;11] | 9 [7;11] | 0.59 [-0.06, 1.25] | 0.51 [-0.28, 1.30] |
| Weight gain velocity (g/kg/d), mean ± SD | 5.7 ± 2.4 | 5.0 ± 2.1 | -0.91 [-1.38, -0.44] | -0.65 [-1.32, 0.01] |
| MUAC gain velocity (mm/w), mean ± SD | 2.3 ± 0.7 | 2.1 ± 0.6 | -0.20 [-0.35, -0.06] | -0.15 [-0.33, 0.03] |
| Number of treatment visits, median [IQR] | 6 [5;8] | 6 [5;8] | -0.27 [-0.84, 0.31] | -0.23 [-0.78, 0.32] |
| Number of missed visits, median [IQR] | 0 [0;0] | 0 [0;0] | -0.16 [-0.33, 0.01] | -0.17 [-0.37, 0.02] |
| Amount of RUTF per child, median [IQR] | 98 [84;126] | 91 [77;112] | -9.13 [-15.31, -2.94] | -8.68 [-15.12, -2.24] |
| **Recovered only** |  |  |  |  |
| Duration of treatment (weeks), median [IQR] | 9 [7;11] | 9 [8;11] | 0.75 [0.26, 1.24] | 0.73 [0.06, 1.40] |
| Weight gain velocity (g/kg/d), mean ± SD | 5.7 ± 2.4 | 5.05 ± 2.07 | -0.88 [-1.35, -0.41] | -0.61 [-1.29, 0.07] |
| MUAC gain velocity (mm/w), mean ± SD | 2.3 ± 0.7 | 2.12 ± 0.59 | -0.20 [-0.34, -0.06] | -0.15 [-0.33, 0.03] |
| Amount of RUTF per child, median [IQR] | 98 [84;126] | 91 [77;112] | -7.97 [-13.10, -2.84] | -6.04 [-11.40, -0.69] |
| <sup>a</sup> adjusted for sex, age, height, MUAC, weight-for-height z-score, height-for-age z-score, month of admission and visit frequency in MAM phase (weekly or fortnightly). |  |  |  |  |

Abbreviations: MAM, moderate acute malnutrition; MUAC, mid-upper arm circumference; RUTF, ready-to-use therapeutic food; SAM, severe acute malnutrition.

## DISCUSSION

To our knowledge, this is the first study testing the change of visit frequency during MAM treatment from weekly to fortnightly visits and the impact of including a 2-week transition period for children admitted as SAM when transitioning to receive a lower RUTF dose in the MAM phase. We showed that these changes do not affect recovery but do result in differences in other treatment outcomes.

### Visit frequency changes

Our finding that visit frequency does not impact recovery are different from a previous study that studied children treated for SAM and showed that recovery was lower and that cumulative mortality was higher by 3-months post-discharge with monthly visits compared to weekly visits [27]. The reason for the discrepancy between our findings and those by Hitchings et al. (2022) may be due to the greater change in visit frequency (monthly throughout treatment in Hitchings versus weekly during SAM and fortnightly during the MAM phase in our study), context (generally more fragile in northwest Nigeria than southwest of Mali), and study population (SAM are more vulnerable than GAM).

While we did not observe differences in recovery of children by visit frequency, we did observe an increase in the duration of treatment by 2.5 weeks. Up to 1.5 weeks of this increased duration of treatment may be explained by protocol stipulations, namely the requirement to meet recovery criteria at two consecutive visits before discharge and the interval between those visits (see Supplementary Figures for illustrations). The remaining extra 1 week may be attributed to the fact that children following fortnightly visits also had a lower weight gain velocity, suggesting that visit spacing reduces treatment efficacy. Reasons for this could be related to later diagnosis and treatment of other illness episodes occurring during malnutrition treatment or sub-optimal adherence to RUTF intake including potentially increased sharing with other household members. We did not collect information on illness episodes or treatment, or RUTF adherence so we cannot investigate the effect of these potential factors on the longer duration of treatment observed with fortnightly visits. Given the generally short duration of treatment (8 and 4 weeks among children admitted with SAM and MAM, respectively) and the equally low proportion (<1% overall) of children ending up non-recovered by 16 weeks into treatment regardless of visit frequency, longer duration of treatment when implementing fortnightly visits in the MAM may not be a problem in terms of program outcomes in this and similar type of contexts.

From the provider perspective, each treatment visit represents a cost to the health system in terms of health care staff time [28,29]. Thus, spacing out visit frequency may be a way to decrease workload and human resource needs. However, the decreased cost related to potentially fewer visits need to be balanced out with the increase in duration of treatment and the consequent increase in RUTF needs. The 2.5 weeks longer duration of treatment resulted in an average of 23 more RUTF sachets needed to treat a child when following a fortnightly visit frequency in the MAM phase (average of 73 sachets prescribed per child) compared to children following weekly visit frequency throughout treatment (average of 48 sachets of RUTF prescribed per child). This represents a nearly 50% increase in RUTF needs to treat a child. We will be investigating the cost implications of the visit frequency changes in a separate study.

Against our expectations, we did not observe an increase in visit adherence (measured as number of missed visits or proportion defaulting) by children following fortnightly visits when compared to those following weekly visits. Adherence to treatment was generally high with <2% children defaulting and very few missed visits reported. We suspect this is because most children in treatment live relatively close to the treatment sites with >50% living in the village or town where treatment is provided [30]. Also, the continuous support and supervision to the treatment activities is expected to increase adherence [31]. In settings with challenging access to care due to distance, insecurity or other reasons, spacing visit frequency may be more relevant to decrease opportunity cost of care-seeking [28] and improve adherence [32].

### SAM to MAM transition period

We found that including a 2-week transition period before switching from a higher (2 sachets/day) to lower (1 sachet/day) RUTF dose for children admitted as SAM once they reach MAM status resulted in similar recovery and duration of treatment but slightly lower weight gain velocity (that was only significant in unadjusted analysis). Previously, RUTF dose reductions have mostly been shown not to result in decreased weight gain velocity despite greater RUTF dose reductions being tested [9,10,13,33] with the exception of 2 studies where the study populations were not comparable at baseline and a reduction in weight gain was observed for children treated with reduced RUTF dosages [11,34]. Whether a lower weight gain velocity should be of concern remains debated with a need to balance short- and long-term outcomes [35]. Rapid weight gain helps children out of the high risk “zone” of low anthropometry [36] but very high (13 g/kg/d) weight gains during inpatient care have been shown to be associated with adult adiposity [37]. In outpatient care weight gain velocities practically never exceed 10 g/kg/d with a mean of 3.9 g/kg/d observed across 60 programs [21]. Given the relatively high weight gain velocity among children with SAM in our study population (on average >5g/kg/d in both groups), the small negative impact on weight gain velocity of the immediate transition might not be of concern in this setting.

We observed a (statistically insignificant) trend for a 3%-point increase in the proportion of children admitted as SAM and who after having reached MAM criteria regressed back to SAM as a result of immediate transition to lower (1 sachet/d) RUTF dose. This suggests that some children with SAM may require a higher than 500kcal/d dosing of RUTF beyond MUAC≥115mm to ensure they continue steady progress towards recovery. Taken together with the relatively small 10 sachet reduction in the amount of RUTF needed to treat a child resulting from immediate transition from SAM to MAM phase from a total of 105 sachets needed, it seems the immediate transition may not be worth the risk. Ultimately, the choice for including or not the transition period may also depend on feasibility for health workers to apply such a transition.

### Strengths and weaknesses

This study has several strengths including the high completeness and quality of data as evidenced by low number (n=20) of patients with unknown treatment outcomes. Additionally, the study was based on a routine program which strengthens our confidence on external validity of findings. The study also suffers from several limitations. First, the imperfect implementation of the interventions in some health areas due to limited support and supervision of routine staff responsible for making the changes in treatment practices resulted in high proportion of exclusions from the PP analysis for some arms. Given the consistency between the results from the ITT and PP analyses we however expect this limitation not to change the results. Second, while routine program implementation may be a strength in terms of external validity, it did limit the availability of additional data at baseline. For example, we cannot speak to potential underlying differences between health areas potentially due to inherent differences in challenges for caregivers to access treatment. However, we believe that the number of clusters included per intervention for the analysis of differences (n=19-20 clusters) helps balance most baseline differences.

### Generalizability

These results are probably generalizable to context where acute malnutrition treatment is relatively accessible including thanks to treatment being provided by CHWs and where distances to treatment are relatively short [30]. We did not see an impact of spacing out visit frequency from weekly to fortnightly on defaulting but this could be different in a context where distance and access plays a larger role in explaining programmatic outcomes. However, the results on the impact of these changes on duration of treatment, RUTF consumption and weight and MUAC gain velocity are expected to stay robust to context and thus speak in favor of weekly visit frequency throughout treatment and inclusion of a 2-week SAM to MAM transition period before reducing RUTF dose to ensure high weight gain velocity.

## Conclusion

This study investigated the effect of changes in visit frequency and SAM to MAM transition on the effectiveness of acute malnutrition treatment in Mali. We showed that spacing visit frequency in the MAM phase of treatment from weekly to fortnightly did not affect recovery but did increase duration of treatment and RUTF consumption and decrease weight gain velocity. We also showed that among children admitted as SAM immediate transition to a lower RUTF dose once they reach MAM criteria when compared to applying a 2-week transition did not affect recovery rates nor duration of treatment but led to a small decrease in weight gain velocity and possibly to an increase in the number of children regressing back to SAM. Based on these findings, we recommend applying weekly visits throughout treatment where feasible and including a 2-week transition period before reducing the RUTF dose for children admitted with SAM once they reach MAM criteria.

## Data Availability

Data can be accessed freely from Zenodo: https://doi.org/10.5281/zenodo.17716792

## Acknowledgements

We would like to thank Bouaré Cissé for managing the field implementation of the study, Alhousseyni Haidara for managing data during field implementation, and Bareye Ouologuem for technical support and supervision during implementation.

## Author contributions

SK conceptualized and designed the study, wrote the original draft and reviewed and finalized the manuscript. ZT conducted the formal analysis. JB acquired the funding. INC and CTO set up and supervised the data collection and managed the project on the ground. All authors reviewed and provided critical feedback to the manuscript.

